# From GWAS to drug: A framework for drug candidate prioritisation using a gene expression signature matching approach

**DOI:** 10.64898/2026.04.22.26349470

**Authors:** Solal Chauquet, Jiayue-Clara Jiang, Lauren F. Barker, Zoe L. Hunter, Gagandeep Singh, Naomi R. Wray, Allan F. McRae, Sonia Shah

## Abstract

Drug targets supported by human genetic evidence have significantly higher approval rates, making genome-wide association studies a valuable resource for drug candidate prioritisation. Transcriptome-wide association study signature-matching is an emerging *in silico* approach that integrates GWAS data with expression quantitative trait loci to generate a disease gene expression signature, which is then compared against drug perturbation databases such as the Connectivity Map. Despite recent adoption, there is no consensus on optimal methodology. Here, we systematically benchmark key parameters, including TWAS method, eQTL tissue model, similarity metric, gene set size, and CMap cell line, using LDL cholesterol, familial combined hyperlipidemia, and asthma as proof-of-concept traits. We demonstrate that while TWAS signature-matching can successfully prioritise known first-line treatments, performance is highly sensitive to parameter choice; for instance, the selection of the cell line used for drug signatures alone can dramatically alter drug prioritisation. Based on these findings, we propose a best-practice framework for robust, genetically-informed drug prioritisation using TWAS signature-matching.

## Introduction

The attrition rate for drugs entering Phase I trials is reported to be around 90%^1,2^. This high failure rate is largely due to preclinical models inadequately predicting efficacy and safety in humans^3^. With the availability of large genomic datasets, human genetic evidence is increasingly being leveraged to inform drug development^4^. Unlike other molecular markers (e.g. RNA and protein) that can often be biased by reverse causality, genetics provides evidence of the causal role of genes in disease^4^. Recent studies have demonstrated that drug targets with human genetic evidence have a more than 2.6-fold higher likelihood of reaching market^5,6^. This association between genetic evidence and increased success rate is largely unaffected by effect size or allele frequency^4^, making the abundance of well-powered genome-wide association studies (GWAS) a valuable resource for informing drug candidate prioritisation.

Drug target Mendelian randomisation (MR) is a commonly used *in silico* approach that utilises GWAS data for drug candidate prioritisation^7^. Several frameworks for robust drug target MR analyses have been proposed^8,9^. However, this approach focuses on single genes as opposed to biological pathways, and can only be used to investigate drugs or compounds where the mechanisms of action(s) (MoA i.e. target gene(s)) are known. For many approved drugs and experimental compounds, the full repertoire of their targets remains unknown, potentially leading to missed therapeutic opportunities. At the same time, several large novel compound catalogues exist providing rich resources for drug discovery. However, as the MoAs for many of these compounds are unknown, their potential therapeutic effects cannot be investigated using an MR approach.

An alternative approach that can overcome the limitations of drug target MR is gene expression signature matching^10^. Here, a disease-associated gene expression signature is identified and compared to a database of drug gene expression signatures. A drug or compound whose gene expression signature is strongly negatively correlated to the disease signature is considered to have strong potential to ‘reverse’ disease-associated gene expression changes and, therefore, a good candidate for further investigation. Connectivity Map (CMap) and L1000^10,11^ are databases of compound signatures generated using cell-based exposure studies that have been extensively utilised for drug discovery via the gene expression signature matching approach. For example, Liu et al^12^ used a gene expression signature for obesity derived from mouse models to query CMap, identifying Celastrol, a compound derived from *Tripterygium wilfordii*, as a drug candidate for obesity. Celasterol was subsequently shown to be a leptin-sensitiser, regulating appetite and energy expenditure, demonstrating the use of signature matching to prioritise compounds even when the MoA is unknown.

However, disease gene expression signatures derived from animal models may not always be reflective of disease in humans^13^, while identifying disease signatures by comparing gene expression in human cases and controls may not be feasible in disease-relevant tissues, and likely reflect both causal and consequential changes of disease. An alternative approach to identifying a disease gene expression signature that likely reflects causal expression changes is through the integration of GWAS results with tissue-specific expression quantitative trait loci (eQTL). This method, known as a transcriptome-wide association study (TWAS), imputes a genetically-predicted gene expression signature for disease, which can then be used to query compound signature databases like CMap to prioritise drug candidates whose expression signature displays a strong negative correlation with the disease signature (Fig. 1). This requires neither knowledge of drug MoA, nor an understanding of the causal disease pathways, providing a hypothesis-free approach to drug candidate prioritisation informed by human genetics.

**Figure 1.**
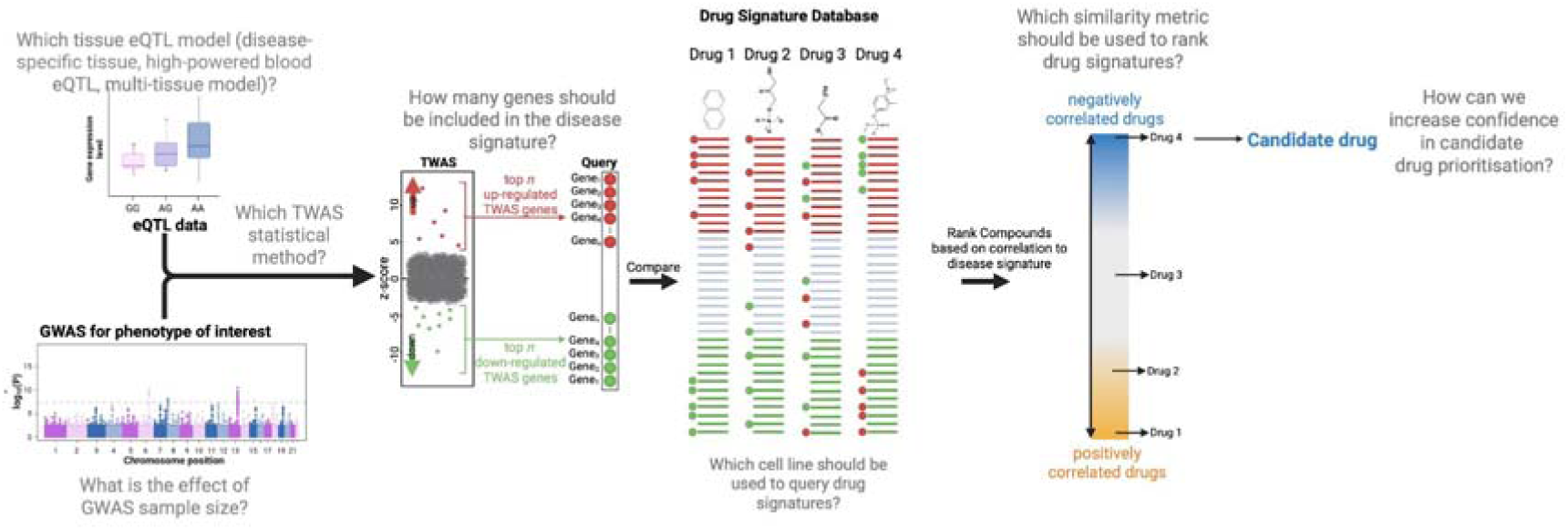
Schematic of the TWAS-based drug repositioning pipeline and benchmarking strategy. a. Overview of the drug repositioning framework. Briefly, summary statistics from a disease-specific GWAS are integrated with relevant eQTL data to perform a Transcriptome-Wide Association Study (TWAS). The resulting associations serve as a disease-specific gene expression signature. A specific subset of these genes is compared against a database of drug gene expression signatures. Candidate therapeutics are identified by their ability to reverse the disease signature; thus, a strong inverse (negative) correlation suggests therapeutic potential. Variables that may influence compound shortlisting are highlighted, with specific research questions addressed in this study indicated in grey.

Though TWAS signature-matching is increasingly being used^14–23^, there is a lack of consensus on methodology and sufficient justification for the choice of key variables. A summary of the divergent approaches to TWAS signature-matching is presented below and in Supplementary Table 1:

1. **TWAS methodology and eQTL models**: Different methods exist to generate TWAS signatures, with FUSION^24^ and sPrediXcan^25^ being the most common. These methods can be applied to tissue-specific or multi-tissue eQTL models. Given the overall shared nature of eQTLs across tissues^26,27^, pooling eQTL information from multiple tissues can increase the power of TWAS. Multi-tissue models or whole blood-based QTL models are often used when the relevant disease tissue is either unknown or lacks well-powered eQTL data. However, the impact on drug prioritisation when selecting a well-powered multi-tissue or blood eQTL model over a lesser-powered, but disease-relevant tissue eQTL model has not been investigated.
2. **Number of TWAS genes used to query CMap**: While some studies use only genes that had a statistically significant p-value (Bonferroni or FDR) in TWAS analysis^14,16^, others have used an equal, but arbitrary number of the most significant up- and down-regulated TWAS genes^19,21^ without providing strong justification for the choice of the TWAS gene set size.
3. **Drug perturbation cell line**: For most studies, the cell line from which drug expression signatures were extracted and compared was not explicitly stated (Supplementary Table 1). Some studies rank compounds based on the similarity of the TWAS signature with drug gene expression signatures profiled across nine core cell lines in CMap which are derived from different tissues^11^. How this impacts the final drug ranking has not been investigated.
4. **Similarity metric**: Most published studies use the normalised connectivity score (NCS) as well as the Tau score, two metrics developed by the CMap authors and implemented in the online CMap/iLINCS portal^11,28^, to compare their TWAS signature to CMap drug signatures. Some studies have used Spearman correlation to rank compounds. However, a direct comparison of drug prioritisation using these different similarity metrics has not previously been presented.

This highlights the need for systematic evaluation of the performance of the TWAS signature-matching approach (Fig. 1). To address this knowledge gap, we used LDL-C, hyperlipidemia and asthma as proof-of-concept traits to evaluate the impact of the different variables described above on prioritisation of known drugs for these traits. The choice of quantitative and disease proof-of-concept traits was based on the availability of well-powered GWAS data^29–32^ (Supplementary Table 2) and first-in-line treatments for these traits being well-profiled across different cell lines in CMap. HMGCR inhibitors (statins) inhibit cholesterol production in the liver and are used for treating high cholesterol and metabolic syndromes such as familial combined hyperlipidemia^31^. Corticosteroid agonists have potent anti-inflammatory effects and are first-line treatments for asthma^33^. As the full range of drug effects is unknown, it is impossible to determine if a prioritised drug is a false positive without extensive experimental validation. Therefore, our evaluation focuses on the ranking of first-line treatments (i.e. high ranking of true positives), to inform the most optimal parameters for drug prioritisation. Though published studies focus on ranking of individual compounds, if a particular MoA is effective against a disease, one would expect compounds that share the same MoA to rank highly when using the TWAS signature-matching approach. Therefore, enrichment of a drug class provides a more robust way of evaluating drug prioritisation.

To benchmark the performance of TWAS signature-matching, we first performed a TWAS, followed by a comparison of TWAS signatures of varying gene set sizes to individual CMap compound signatures. The TWAS gene sets included 144 sets with varying numbers of up-and down-regulated TWAS genes, ranging from a total of 10 (5 up- and 5 down-regulated) to 120 genes (60 up- and 60 down-regulated). The maximum number of genes included within the query was selected based on the median number of differentially expressed genes per compound in CMap (see Supplementary Note). Individual compounds were then ranked by similarity and used to perform a drug class enrichment analysis using the well-known GSEA algorithm^34^ which provides a normalised enrichment score (NES). A strong negative NES indicates that compounds within a given drug class are enriched in the negative tail of the similarity ranking, meaning that members of that class tend to negatively correlate with the TWAS signature more often than would be expected by chance. In the case of a true positive drug class, the expectation would be consistent (negative) enrichment of the drug class across multiple TWAS gene sets. Therefore, final drug class ranking was based on the mean NES across all tested TWAS gene sets. For reproducibility, we used a locally re-processed version of CMap (see Supplementary Note) containing 221,549 unique drug perturbation consensus signatures based on a minimum of 3 replicates.

By clarifying the strengths, limitations, and best-use scenarios, we provide a set of recommendations to optimise TWAS signature-matching analyses for robust prioritisation of drug candidates.

## Results

### Drug prioritisation for LDL-C

#### Impact of similarity metric and TWAS gene set query size

Since the liver is the most biologically relevant tissue for cholesterol production, we generated TWAS signatures using the European GWAS data for LDL-C (1,320,016 individuals) and sPrediXcan with a liver eQTL model based on Genotype Tissue Expression (GTEx) liver eQTL data (Supplementary Tables 4). LDL-C TWAS gene sets of varying size were compared to all drug signatures in CMap profiled in HEPG2 cells, a liver-derived cancer cell line. Compounds were ranked based on either Spearman correlation or NCS and used to calculate a drug class enrichment using GSEA. Drug classes were then ranked by mean NES (drug classes with the most negative NES ranked highest).

When drug-trait signature similarity was calculated using Spearman correlation, HMGCR inhibitors were consistently significantly enriched across almost every tested TWAS gene set (Fig. 2a-left) and ranked first among all tested drug classes (mean NES±SD=-1.76±0.15 p=1.46 x 10^−02^; Supplementary Table 5), demonstrating the ability to prioritise first-line treatments using the TWAS signature-matching approach. However, when using the NCS metric to rank compounds (Fig. 2a-right), HMGCR inhibitors were ranked fourth and were not significantly enriched (mean NES=-1.26±0.25, p=2.0 x 10^−01^; Supplementary Table 6).

**Figure 2.**
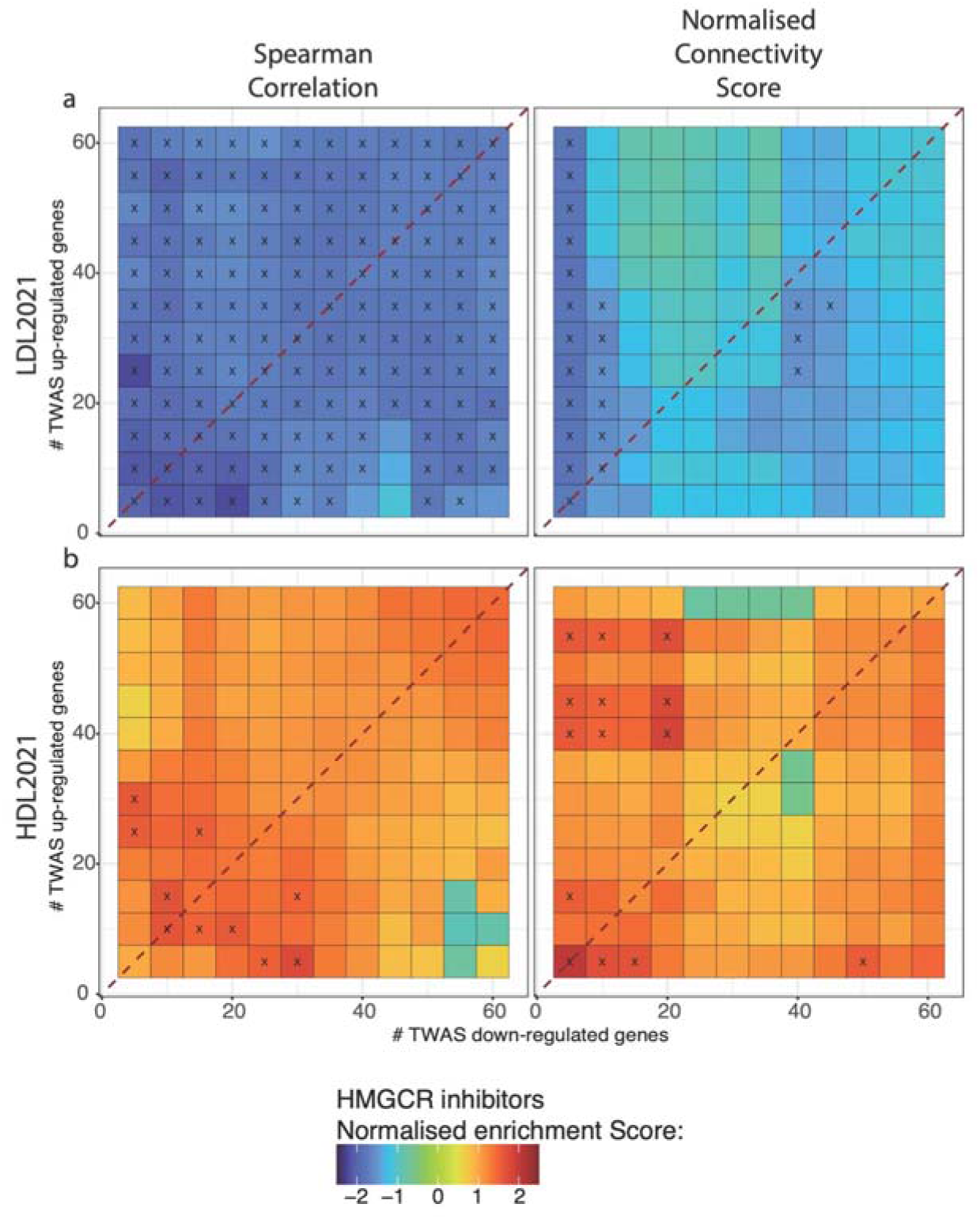
Impact of similarity metric on enrichment of HMGCR inhibitors for LDL-C and HDL-C TWAS signature. Similarity between drug gene expression signatures and TWAS-derived signatures for (a) LDL-C and (b) HDL-C was evaluated across varying gene set sizes. Drug similarity was calculated using either Spearman correlation (left panels) or the Normalized Connectivity Score (NCS) (right panels) for compounds screened in the HEPG2 liver cell line. For each gene set combination, HMGCR inhibitor enrichment was quantified using the GSEA algorithm. The x and y axes represent the number of down-regulated and up-regulated TWAS genes (ranging from 5 to 60) included in the similarity calculation. The red dotted line indicates queries with a symmetric distribution of up- and down-regulated genes. Black crosses indicate significant enrichment defined as an absolute NES > 1.5.

Interestingly, when restricting the LDL-C signature to a single gene set comprising of only statistically significant TWAS genes (103 up- and 100 down-regulated genes; Supplementary Figure 1), although HMGCR inhibitors were overall negatively correlated with the LDL-C signature, the enrichment was no longer statistically significant with either similarity metric (Spearman: mean NES=-1.33±0.30, p=1.63 x 10^−01^, ranked 8th; NCS: NES=-0.97±0.65, p=4.89 x 10^−01^, ranked 15th), demonstrating the value in testing multiple gene set sizes.

Although not one of their main effects, statins have been shown to modestly increase HDL-C^35^. We therefore also tested a TWAS for high HDL-C^29^ (Supplementary Table 7). Though HMGCR inhibitors were positively correlated using both metrics (Fig. 2b), they were not significantly enriched using either Spearman correlation (mean NES±SD of 1.13±0.40 and a p-value of 5.60 x 10^−01^, Supplementary Table 8) or NCS (1.07±0.45 and 5.40 x 10^−01^; Supplementary Tables 9).Overall, these results for LDL-C (strong negative correlation) and HDL-C (weak positive correlation) are consistent with the known strong LDL-C-lowering effect and weak HDL-C-increasing effect of HMGCR inhibitors.

Given that Spearman correlation led to more consistent and strongest prioritisation of HMGCR-inhibitors compared to NCS, all subsequent analyses used Spearman correlation for drug-trait comparison.

#### Impact of GWAS power

Next, we conducted a sensitivity analysis using a less-powered GWAS of LDL-C published in 2013^30^ (Effective sample size - Neff = 89,872, compared to Neff = 647,692 for the 2021 LDL-C GWAS). Both studies identified a large number of independent, genome-wide significant loci, with 58 in the 2013 GWAS and 403 in the 2021 GWAS. While the difference in sample size is stark, the genetic correlation between the two studies was close to 1 (rg±se=0.99±0.02, Supplementary Table 10). TWAS signatures from the two GWAS were also strongly correlated (r=0.75, p<2.20 x 10^−16^, Supplementary Figure 3). When performing a signature-matching approach using the 2013 derived TWAS (Supplementary Table 11) and HEPG2 cells, enrichment of HMGCR inhibitors diminished slightly, but were still the second most negatively enriched drug class with a mean NES of −1.62±0.23, (p=4.64 x 10^−02^) (Supplementary Figure 2, Supplementary table 12).

#### Impact of TWAS method

Next, using the liver eQTL model and HEPG2 drug signatures, we compared two common TWAS methods, sPrediXcan and FUSION (Fig. 3, Supplementary Table 13). Despite TWAS signatures from the two methods showing strong positive correlation (r=0.90, p<2.20 x 10^−16^, Supplementary Figure 3), HMGCR inhibitors were only significantly enriched and ranked first when using sPrediXcan (−1.76±0.15 p=1.46 x 10^−02^). The FUSION TWAS led to a lower, non-significant enrichment (mean NES±SD = −1.14±0.17, p=3.7 x 10^−01^, Supplementary Table 14), with HMGCR inhibitors ranked fifteenth.

**Figure 3.**
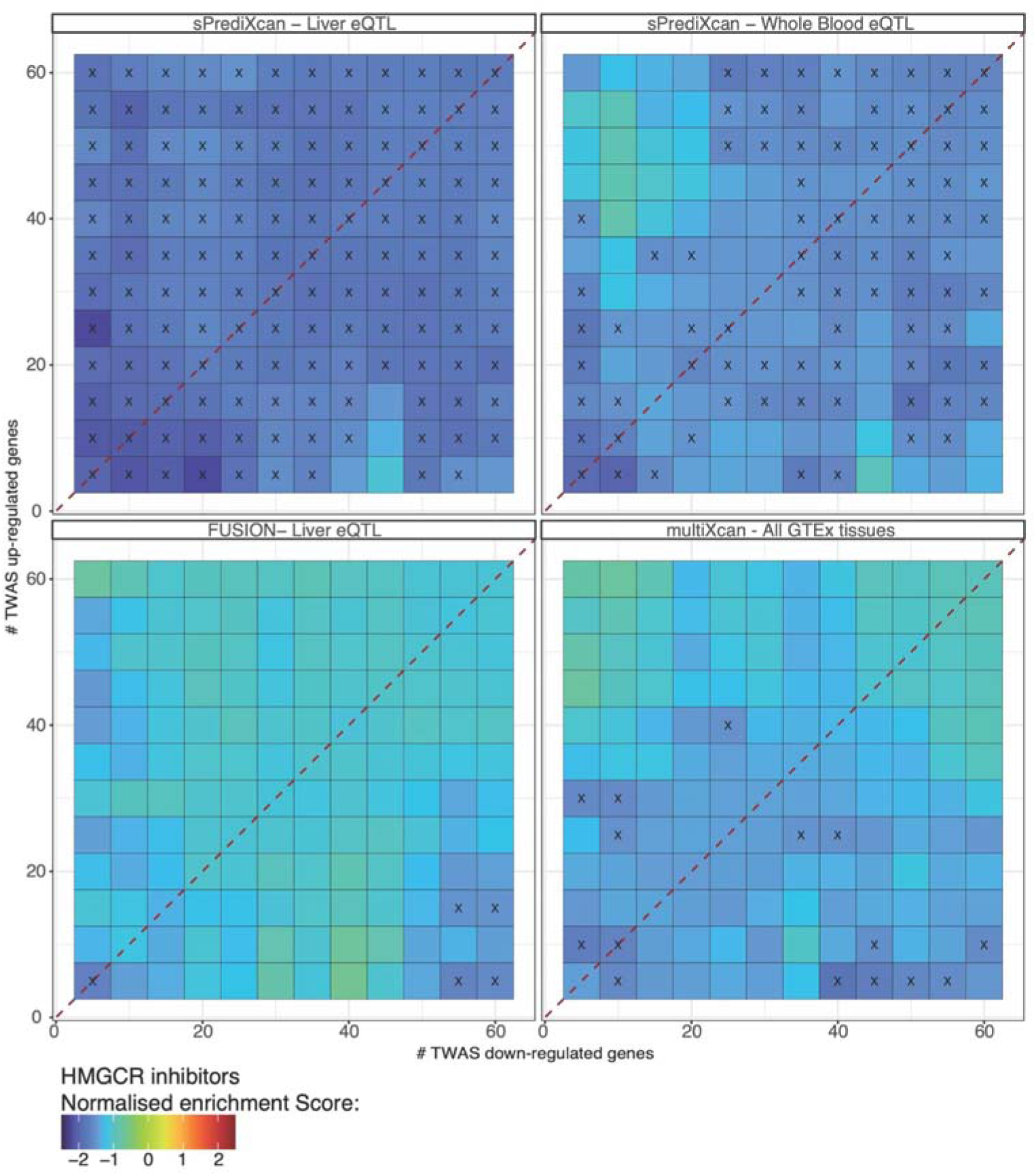
Impact of TWAS method and eQTL model on enrichment of HMGCR inhibitors for LDL-C. Similarity between drug perturbation profiles and TWAS-derived signatures for LDL-C was evaluated across four different methods: (i) sPrediXcan liver eQTL, (ii) sPrediXcan Whole Blood eQTL, (iii) FUSION liver eQTL and (iv) sMultiXcan using all GTEx tissues. Drug similarity was calculated using Spearman correlation for compounds screened in the HEPG2 cell line across varying geneset sizes. For each TWAS/eQTL combination, HMGCR inhibitor enrichment was performed using the GSEA algorithm. The x-axis represents an increasing number of down-regulated TWAS genes included in the similarity calculation, while the y-axis represents a similar increase in up-regulated genes. The red dotted line indicates queries with a symmetric distribution of up- and down-regulated genes. Black crosses correspond to significant enrichment, defined as an absolute NES > 1.5.

#### Impact of tissue-specific eQTL models for TWAS

The LDL-C TWAS signature based on whole blood eQTL data from 755 blood samples (Supplementary Table 15) was only moderately correlated with the TWAS signature based on liver eQTL data from 226 liver samples (r=0.44, p<2.20 x 10^−16^, Supplementary Figure 3). TWAS based on whole blood eQTL led to a weaker and non-significant negative enrichment of HMGCR inhibitors (Fig. 3, mean NES±SD=-1.49±0.19, p=8.6 x 10^−02^, Supplementary Table 14), which were ranked fifth. Similarly, a LDL-C TWAS signature using a multi-tissue eQTL model (Supplementary Table 16) showed moderate positive correlation with the liver TWAS signature (r=0.49, p<2.20 x 10^−16^, Supplementary Figure 3). However, HMGCR inhibitors were not significantly enriched (Fig. 3, mean NES±SD=-1.28±0.21, p=2.4 x 10^−01^, Supplementary Table 14) and ranked ninth. Altogether, these results suggest that biologically-relevant tissue-specific TWAS signatures perform better than those based on multi-tissue models, despite the latter having greater power for TWAS analysis.

#### Impact of cell line selection

Using the gene signature generated from the LDL-C 2021 dataset using sPrediXcan with a liver eQTL prediction model, we assessed the ranking of HMGCR inhibitors based on drug signatures measured in nine core CMap cell lines: A375 (skin), A549 (lung), HA1E (kidney), HCC515 (lung), HEPG2 (liver), HT29 (colon), MCF7 (breast), PC3 (bone), and VCAP (prostate) (Fig. 4a; Supplementary Table 17).

**Figure 4.**
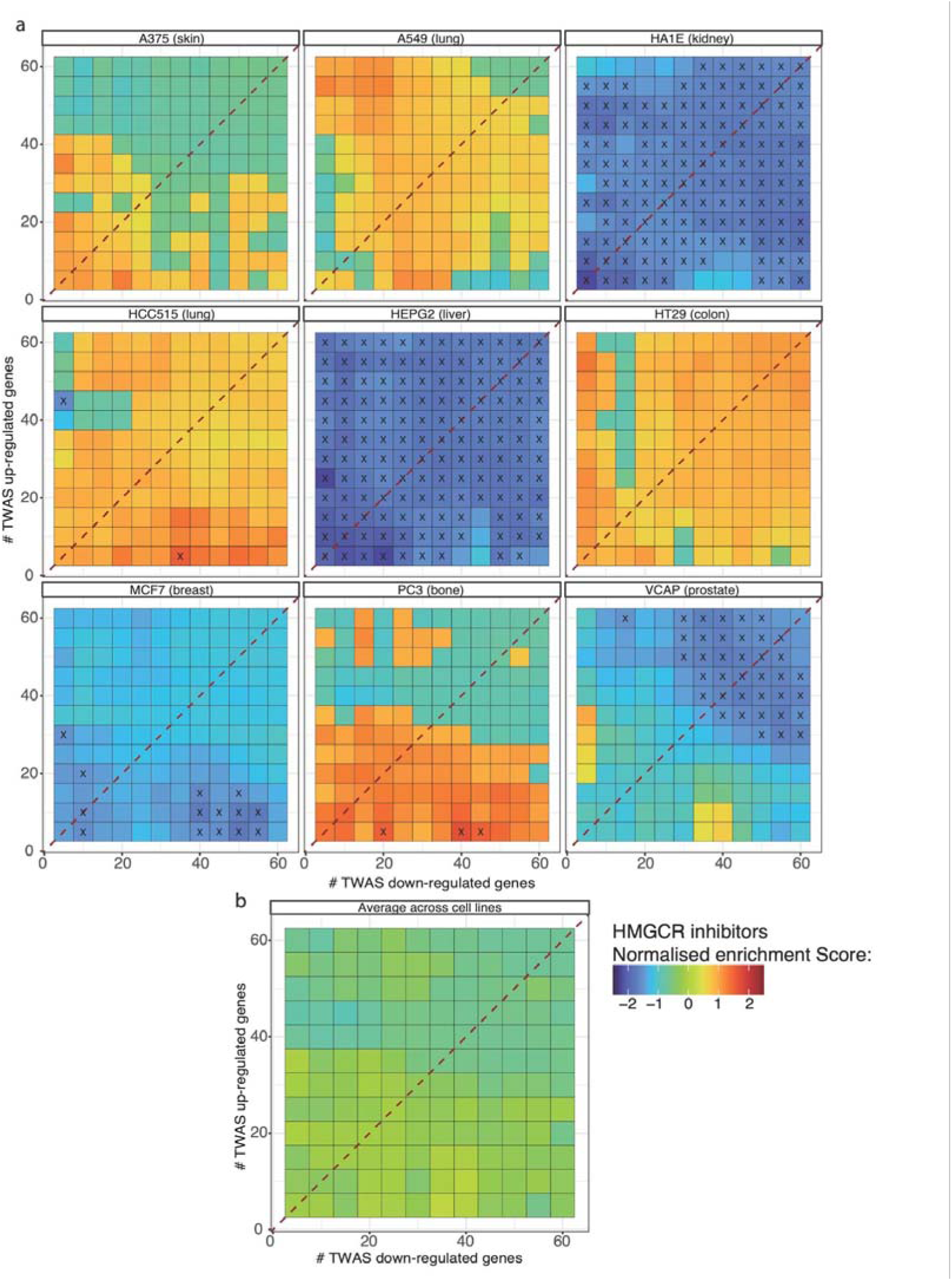
Impact of cell-line specificity on HMGCR inhibitor enrichment signatures for LDL-C. (a) Similarity between drug perturbation profiles and TWAS-derived signatures for LDL-C was evaluated across the nine CMap core cell lines. Drug similarity was calculated using Spearman correlation across varying geneset sizes, with HMGCR inhibitor enrichment quantified via the GSEA algorithm. The x-axis represents an increasing number of down-regulated TWAS genes included in the similarity calculation, while the y-axis represents a similar increase in up-regulated genes.The red dotted line indicates queries with a symmetric distribution of up- and down-regulated genes, and black crosses denote significant enrichment (absolute NES > 1.5). (b) Representation of the average (across cell lines) HMGCR enrichment observed when calculating similarity between TWAS and drugs regardless of cell line of origin, demonstrating no signal and hence the importance of considering only cell lines relevant to the trait

Negative enrichment of HMGCR inhibitors was strongest in the liver-derived HEPG2 cell line (mean NES±SD = −1.76±0.15, p=1.46 x 10^−02^), where they ranked first as a drug class. This was followed by the kidney-derived HA1E (−1.64±0.16, p=4.6 x 10^−02^) where HMGCR inhibitors were ranked 7th. In contrast, HMGCR inhibitor signatures exhibited either weak negative or even positive correlation with the LDL-C TWAS signature in the other cell lines (MCF7 −1.28±0.15; p=2.5 x 10^−01^, rank=30; VCAP −1.16±0.52; p=3.1 x 10^−01^, rank=24; A375:-0.15±0.75, p=9.9 x 10^−01^, rank=139; PC3: 0.28±1.03, p=9.9 x 10^−01^, rank=167; A549: 0.50±0.63, p=9.6 x 10^−01^, rank=148; HCC515: 0.75±0.51; p=8.5 x 10^−01^, rank=134; HT29: 0.75±0.50, p=8.4 x 10^−01^, rank=183), illustrating the substantial variability in drug ranking by choice of cell line. Several published studies use an average across cell lines to rank compounds. However, when aggregating results across all nine cell lines by performing the enrichment using all compounds regardless of cell line, the overall enrichment was non-significant (−0.41±0.22, p=9.8 x 10^−01^; Fig. 4b). This suggests that gene expression responses to HMGCR inhibitors across different cell lines vary substantially despite the target HMGCR protein being ubiquitously expressed in human tissues. This emphasises the need for careful selection of the cell line in which drug signatures have been profiled.

### Drug prioritisation for familial combined hyperlipidemia (FCH)

A TWAS signature for FCH was generated based on a case-control GWAS with 17,485 cases and 331,737 controls, using sPrediXcan with a liver eQTL model, and drug signatures profiled in HEPG2 (Supplementary Table 18). Though a negative enrichment was observed, this was not statistically significant (−1.44±0.27, p=1.58 x 10^−01^, Fig. 5a). HMGCR inhibitors were ranked ninth as a drug class (Supplementary Table 19). While the GWAS of FCH identified 170 genome-wide significant loci, TWAS using a liver eQTL model only identified 17 significant TWAS genes. Restricting the query to include only genesets containing fewer than 20 genes, we obtain an overall significant enrichment of HMGCR (−1.72±0.15, p=3.48 x 10^−02^, rank=7).

**Figure 5.**
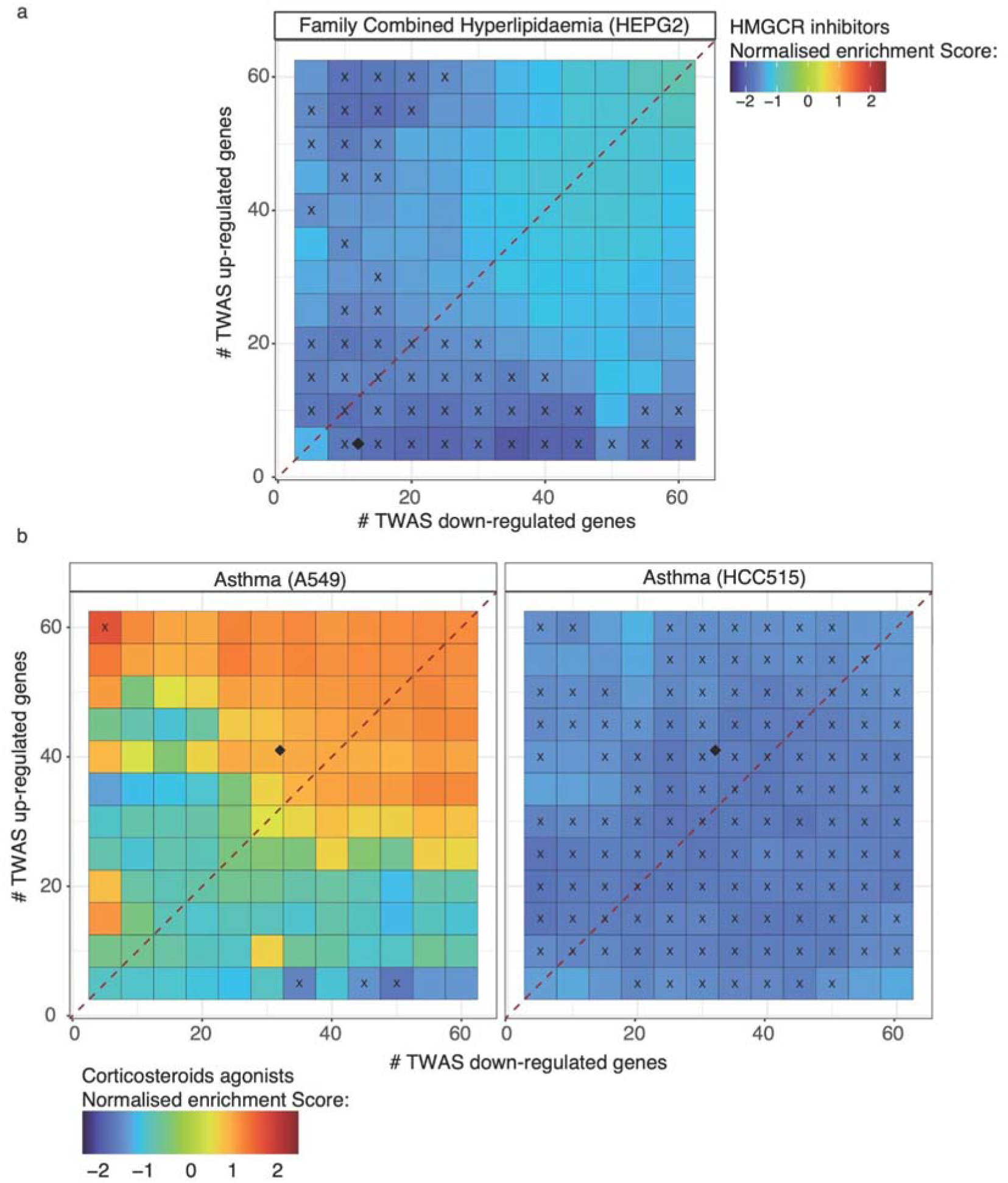
Application of TWAS-repurposing on binary traits: familial combined hyperlipidaemia and asthma. Normalised enrichment score for HMGCR inhibitors for familial combined hyperlipidaemia (FCH, a) and for corticosteroid agonists for asthma (b). Drug similarity was calculated across varying geneset sizes using Spearman correlation for compounds screened in the HEPG2 cell line for FCH and either A549 or HCC515 for asthma. Respective drug class enrichments were then calculated using the GSEA algorithm. The x-axis represents an increasing number of down-regulated TWAS genes included in the similarity calculation, while the y-axis represents a similar increase in up-regulated genes. The red dotted line corresponds to a query containing a similar number of up and down regulated genes. Black cross corresponds to a significant enrichment defined as an absolute NES > 1.5. The Black diamond corresponds to the query size when including all significant TWAS genes of the respective TWAS used for repurposing.

### Drug prioritisation for asthma

TWAS signature was generated using sPrediXcan with a lung eQTL model (Supplementary table 20). Drug signatures from two lung cancer tissue-derived cell lines: HCC515, a lymph node derived cell line, and A549, an epithelial cell line were queried (Fig. 5b). Corticosteroids agonists were the second most negatively enriched drug class (−1.64±0.11, p=2.0 x 10^−02^, Supplementary Table 21) when using the HCC515 cell line. However, no enrichment of corticosteroid agonists was observed when using drug signatures profiled in the lung epithelial cell line (0.06±0.99, p=9.9 x 10^−01^, rank=252). Though both immune and epithelial cells are relevant in asthma, GWAS signals for asthma show the strongest colocalisation with DNase I hypersensitive sites in immune cell types and specifically B-cells^36^. The specificity of drug class enrichment in only the lung lymph node-derived cell line is consistent with the anti-inflammatory and immunosuppressive mechanism of action of corticosteroid agonists.

The enrichment scores and ranking of first-line drug classes in all the above analyses is summarised in Table 1.

**Table 1.** Summary of the systematic evaluation of the performance of TWAS signature-matching for drug prioritisation for four proof-of-concept traits.

| GWAS | eQTL model | TWAS method | drug signature cell lines | similarity metric | Performance |  |  |
| --- | --- | --- | --- | --- | --- | --- | --- |
|  |  |  |  |  | mean NES±SD | Enrichment p-value | Ranking of first-line treatment drug class |
| LDL 2021 | Liver | sPrediXcan | HEPG2 | Spearman | -1.76±0.15 | 0.014 | 1 |
| LDL 2013 | Liver | sPrediXcan | HEPG2 | Spearman | -1.62±0.23 | 0.046 | 2 |
| LDL 2021 | Liver | sPrediXcan | HEPG2 | NCS | -1.26±0.25 | 0.2 | 4 |
| LDL 2021 | Blood | sPrediXcan | HEPG2 | Spearman | -1.49±0.19 | 0.086 | 5 |
| LDL 2021 | All GTEx tissues | sPrediXcan | HEPG2 | Spearman | -1.28±0.21 | 0.24 | 9 |
| LDL 2021 | Liver | FUSION | HEPG2 | Spearman | -1.14±0.17 | 0.37 | 15 |
| LDL 2021 | Liver | sPrediXcan | Average of 9 cell lines | Spearman | -0.41±0.22 | 0.98 | 64 |
| FCH | Liver | sPrediXcan | HEPG2 | Spearman | -1.44±0.27 | 0.16 | 9 |
| Asthma | Lung | sPrediXcan | HCC515 | Spearman | -1.64±0.11 | 0.02 | 2 |
| Asthma | Lung | sPrediXcan | A549 | Spearman | 0.06±0.99 | 0.99 | 252 |

## Discussion

Human genetic evidence is increasingly being used to inform drug discovery and repositioning opportunities^4^. TWAS signature-matching is an emerging approach for drug prioritisation informed by human disease genetic associations. However, unlike drug target MR, relatively few studies have used this approach, with most lacking any validation of prioritised drugs, making it challenging to assess the performance of this approach. In addition, the utility of CMap drug signatures for drug discovery or repurposing has also come under recent criticism due to poor drug signature reproducibility, relevance of drug signatures measured in cancer cell lines, and the limited number of case studies with successful repositioning that have made it to market, despite the resource being made available more than a decade ago^37,38^. Despite these limitations, we demonstrate the ability of the TWAS signature matching approach to successfully prioritise known first-line treatments for proof-of-concept traits, identifying it as a valuable approach for genetically-informed drug repositioning. However, we also highlight how the choice of parameters can drastically impact drug prioritisation. Given the lack of methodological consensus, we draw on our observations to propose a ‘best-practice’ framework for the robust identification of promising drug candidates (Fig. 6), as well as a checklist of the key decision points needed for reproducible TWAS-based drug repurposing (Supplementary Fig. 4).

**Figure 6:**
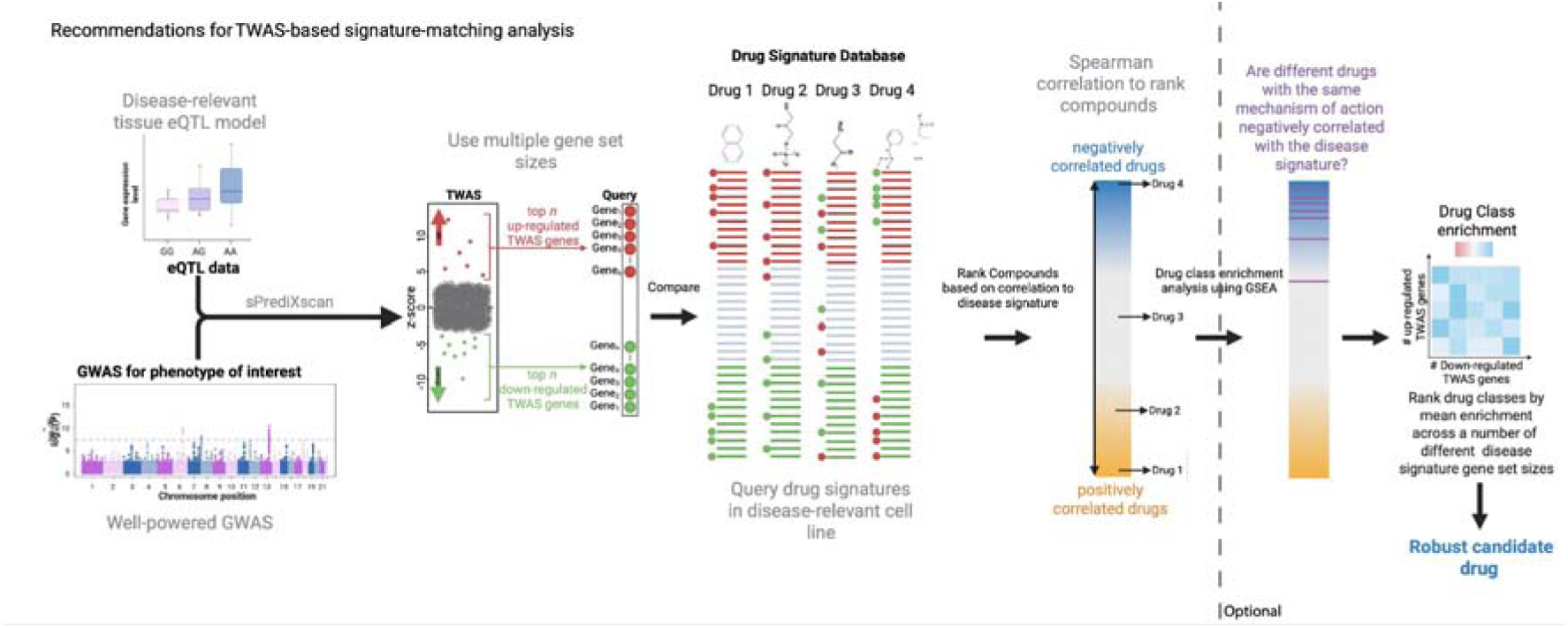
Recommended pipeline for performing TWAS-based drug prioritisation. The figure provides recommendations for TWAS signature-matching analyses for drug prioritisation based on systematic evaluation of this approach to prioritise first-line treatments for three proof-of-concept traits.

Currently, Clue^11^ and iLincs^28^, two online portals, enable drug signature matching with CMap data, primarily using the NCS similarity metric to quantify drug ranking. Using a GWAS of LDL-C, we show that NCS performed poorly compared to Spearman correlation in prioritising HMGCR inhibitors. One potential explanation for the difference in performance is that NCS compares up- and down-regulated genes separately, while Spearman correlation is calculated using both up- and down-regulated genes together, and thus may better capture drug-trait relationships. More recently developed similarity metrics such as XCos^39^, zhangscore^40^, and eXtreme Sum score^41^, display improvement over NCS^42^. However, these methods are not currently widely used. A comprehensive comparison of these different similarity metrics using a similar benchmarking approach could help further inform analyses.

Similarly, several methods for TWAS have been proposed, with FUSION^24^ and sPrediXcan^25^ being the most popular. The main difference between sPrediXcan and FUSION is the handling of SNPs present in the eQTL prediction models but absent in the GWAS of interest. sPrediXcan ignores and removes them from the prediction model while FUSION uses the ImpG-summary algorithm^43^ to impute GWAS missing values and uses them for gene expression prediction. Though TWAS signatures from the two methods showed strong correlation (Supplementary Figure 3), ranking of HMGCR inhibitors was highly variable, with sPrediXcan outperforming FUSION. The potential deleterious impact of imputation on TWAS strength has been raised previously when comparing performance of FUSION and sPrediXcan^44^. Another approach to TWAS methodology is to leverage shared eQTLs across tissues^26,27^ to increase power for predicting a trait gene expression signature. Methods such as sMultiXcan^45^ were developed for this purpose. However, strongest prioritisation of HMGCR inhibitors was observed with a less-powered, liver-specific eQTL model, suggesting that the gain in statistical power for TWAS from multi-tissue models may come at the cost of tissue-specific signatures that are more relevant to disease aetiology and drug action.

Published TWAS signature-matching studies^14–23^, have either used an arbitrary number of genes following TWAS analysis, often equally balanced between the number of up- and down-regulated genes, or all statistically significant TWAS genes. Though the latter seems intuitively more appropriate, HMGCR inhibitor enrichment was not statistically significant when including all TWAS-significant genes (103 up-, 100 down-regulated genes, Supplementary Figure 1). Rather, the strongest enrichment for HMGCR inhibitors was observed when using smaller, unbalanced gene set sizes (ranging from 5 up- and 5 down-regulated genes up to 60). The same behaviour is observed for both FCH and asthma, where drug class enrichment diminished with the inclusion of a large number of genes in the trait signature. This may partly reflect the power of the drug perturbation studies, given most drugs in CMap were only profiled in triplicates, with the median number of differentially expressed genes across all drug signatures being around 120 (61 up-, 59 down-regulated genes, Supplementary Note). Therefore, the inclusion of larger TWAS gene sets may reduce the signal-to-noise ratio when estimating the similarity between drug and trait signatures. Overall, the optimal TWAS signature size may vary depending on the underlying power of the drug perturbation experiment, GWAS and eQTL models, and therefore a single, fixed TWAS gene set size is not recommended. Consistency of drug class enrichment across different TWAS gene set sizes should be used as an indication of the strength of the relationship between drug and disease. Mean enrichment across multiple TWAS gene sets, as well as visual plots, such as the ones presented in this paper, will facilitate robust drug prioritisation.

As mentioned above, statins were only significantly enriched for FCH with smaller TWAS gene set sizes, which may partly reflect GWAS power. However, FCH is characterised not just by elevated plasma concentrations of LDL-C but also apolipoprotein B-100 and triglycerides, as well as low HDL-C. While statins are used as first-line treatment for FCH to lower LDL-C, they are often used in combination with other medications, such as fibrates, which help address the high triglycerides and low HDL-C^46^. Therefore, the lower prioritisation of statins may also reflect their inability to fully address all the underlying disease mechanisms. Interestingly, the most enriched drug class for FCH was Heat Shock Protein (HSP) inhibitors (mean NES −2.12±0.23, p=1.48 x 10^−03^). Though not used as treatment for FCH, HSP90 inhibitors are currently being investigated as a potential treatment for lipid disorders^47,48^, with some evidence to suggest they reduce both cholesterol and triglyceride levels^49^.

HMGCR inhibitors showed the strongest enrichment using drug signatures profiled in biologically relevant cell lines. Given the observed specificity of the HMGCR enrichment across the 9 CMap core cell lines, averaging compound ranking across cell lines, though common in published studies, is not recommended. Furthermore, results on prioritisation of corticosteroid agonists highlight the importance of cell type-specific drug signatures. Asthma is an obstructive inflammatory disease in the lower airways^50^, and immune cells such as lymphocytes and eosinophils play a key role in asthma’s pathophysiology^51^. Despite selecting two lung cell lines for drug signature comparison, a significant enrichment of corticosteroids was only observed in the lymph node derived cell line. Therefore, querying drug signatures profiled in cell lines relevant to disease aetiology is more likely to capture disease-drug relationships. Tissue and cell-type enrichment analysis of GWAS summary data using methods such as MAGMA^52^ can help inform decisions around the choice of eQTL tissue model and drug signature cell line.

Finally, though drug ranking and prioritisation can be performed at an individual compound level, overall ranking of a drug class based on the ranking of multiple, individual compounds with the same MoA (or other information such as indication, biological pathway, etc.) provides greater confidence in results and reduces the possibility of false positives. Identification of a promising MoA can also be followed by drug target MR to provide orthogonal evidence.

In conclusion, we have demonstrated TWAS signature-matching as a promising approach for genetically-informed drug prioritisation. However, careful consideration must be given in its application to ensure translatable outcomes. A summary of these recommendations is presented in Fig. 6.

## Methods

### TWAS-based drug repositioning benchmarking

#### GWAS summary statistics

All GWAS included in this paper are publicly available and details are provided in *Supplementary Table 2*.

#### TWAS analysis

TWAS were performed using three different methods, FUSION^24^, sPrediXcan^25^ and sMultiXcan^45^, since these are the methods that have been applied in published TWAS signature matching studies. TWAS analysis consists broadly of two steps. First the training of models that predict gene expression values from genotypes using data where both genotype and gene expression is measured. Second, the application of those models to GWAS summary data to generate a disease associated genetically predicted gene expression signature. GTExV8^53^ eQTL prediction models were downloaded for sPrediXcan at the following link: https://predictdb.org/post/2021/07/21/gtex-v8-models-on-eqtl-and-sqtl/ and for FUSION here: http://gusevlab.org/projects/fusion/#gtex-v8-multi-tissue-expression. Respective eQTL models were then used with sPrediXcan and FUSION to perform tissue-specific TWAS, and with sMultiXcan to conduct a multi-tissue TWAS for the traits of interest. Regardless of the method, the output from TWAS includes a signed z-score value corresponding to the association between the gene and disease of interest, as well as a p-value indicating the strength of the association. Selection of up- and down-regulated genes for drug signature-matching was based on z-score ranking.

##### TWAS signature gene set size

To evaluate the impact of the number of genes included in the TWAS signature on performance, we systematically varied the number of up- and down-regulated TWAS genes used to query the CMap drug signatures. For each condition, we constructed 144 TWAS gene sets, based on pairwise combinations of a different number of up- and down-regulated genes, ranging from 5 to 60 genes, increasing the number of genes in the up- and down-regulated sets in increments of 5. The similarity between each TWAS gene set and drug signature was then calculated as described below.

#### Similarity metrics

Similarity between TWAS signature gene sets and drug signatures was calculated using either Spearman correlation or the Normalised Connectivity Score (NCS). NCS is defined in the original CMap study^10,11^ and is calculated as follows:

For each query ***q***, consisting of a set of genes genetically predicted by TWAS, genes are partitioned into two subsets: ***q_up_***, containing TWAS genes that are up-regulated (i.e. have a positive TWAS z-score value), and, ***q_down_*** containing genes predicted to be down-regulated (i.e. have a negative TWAS z-score). First, a weighted connectivity score (WTCS) representing a non-parametric similarity metric based on a weighted Kolmogorov-Smirnov like statistic is calculated. This statistic is derived from the GSEA paper^34^. For a query ***q*** and a drug of interest ***d***, we calculate respectively ***ES_up_*** and ***ES_down_***, for the up- and down-regulated gene sets. The WTCS is then calculated as follows:

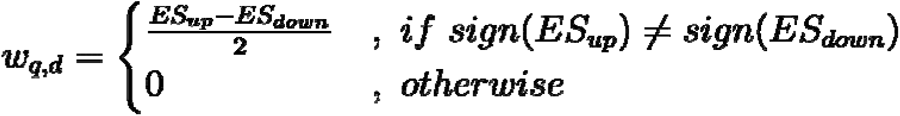

To allow for comparison across different cell types ***c***, the WTCS is then normalised to give the NCS. Given a vector of WTCS values ***w*** resulting from a single query, we normalise the value as follow:

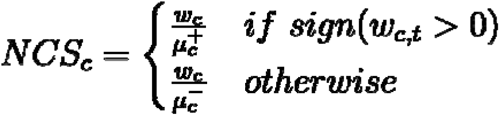

where ***μ^+^*** and ***μ^−^*** are the signed means of the raw weighted connectivity scores evaluated separately.

The NCS ranges from −1 to 1, where a score of 1 indicates that a compound has the strongest positive correlation with the TWAS signature among all tested compounds, and a score of −1 indicates the most negative correlation with the TWAS signature.

Final ranking of CMap compounds was based on the mean similarity metric (either Spearman correlation or NCS) across the 144 TWAS gene sets.

#### Mechanism of action enrichment

Drug class enrichment analysis is then performed based on MoA information provided by CMap using the Gene Set Enrichment Analysis (GSEA) algorithm^34^. GSEA computes a weighted Kolmogorov–Smirnov like statistic to assess whether a predefined group (in this case, different compounds sharing a common MoA) is overrepresented at the extremes of a ranked list. A negative NES indicates that compounds within a given drug class are enriched in the negative tail of the similarity ranking, while a positive NES indicates enrichment among positively correlated compounds. GSEA was restricted to drug classes that contained at least 3 member compounds with gene expression signatures in CMap. Drug class enrichment analysis was conducted using the *fgsea* R package^54^ for each of the 144 TWAS gene sets. Final drug class ranking was based on the mean NES for a drug class across the 144 gene sets. The significance of the mean NES was calculated by comparing the mean value to the empirical distribution obtained from all tested compounds.

#### Code availability

All code used to generate this paper can be found on the following public GitHub repository: https://github.com/SolalC/DrugRepurposing

## Data Availability

All data produced in the present work are already available online.
All code used to generate the results are available on GitHub.
Results from analysis are all available in full in the supplementary tables.

## Acknowledgement

This work was supported by a National Health and Medical Research Council (NHMRC) IDEAS grant (2000637). SS was supported by the National Heart Foundation Future Leader Fellowship (105638).

## Supplementary Figure

**Supplementary Figure 1:**
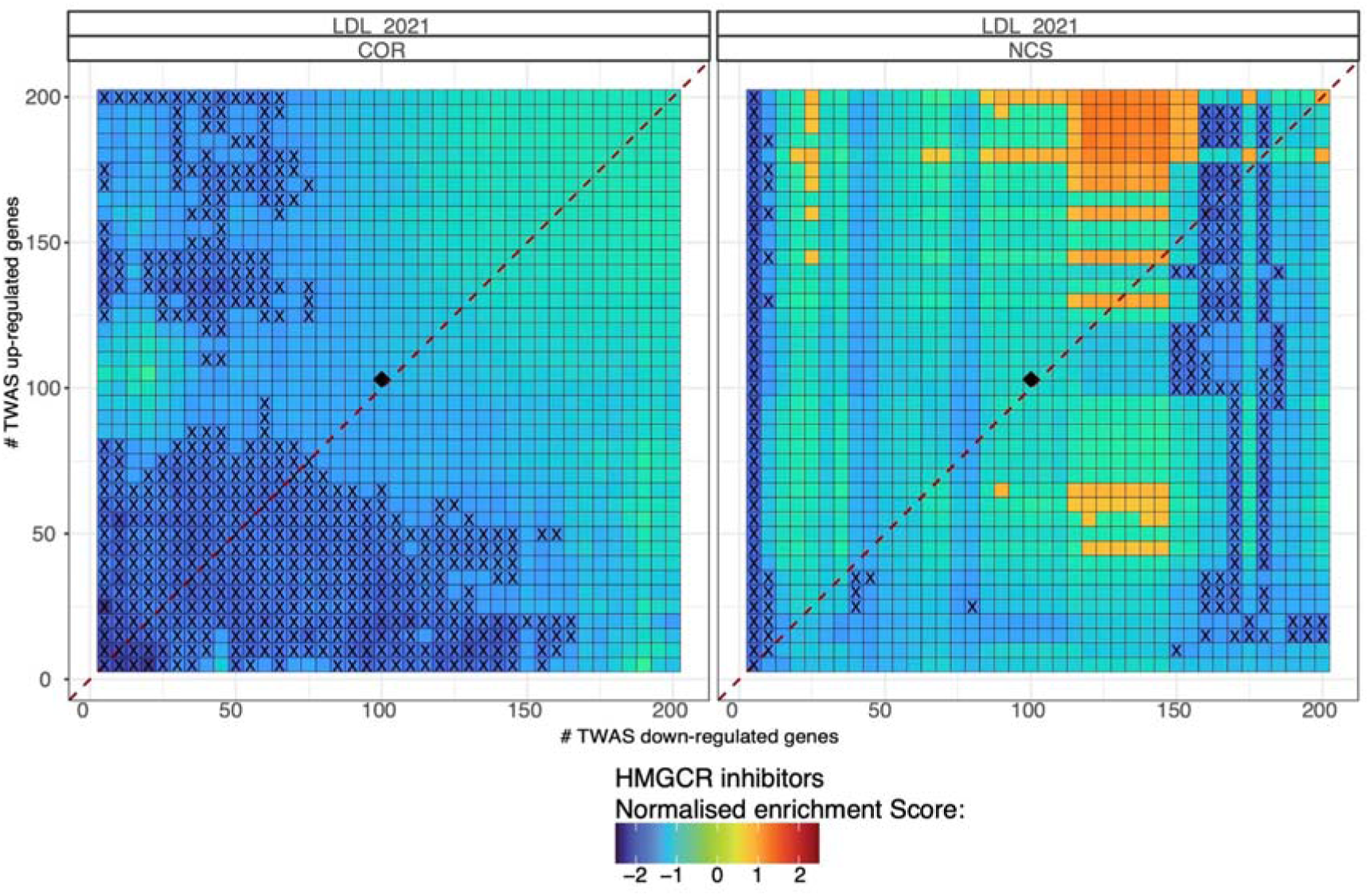
TWAS-based drug repositioning of HMGCR inhibitors for LDL in HEPG2 cell line using genesets containing up to 200 up and 200 down-regulated genes. Normalised enrichment score for HMGCR inhibitors. Individual drug similarity to TWAS signature of LDL-C was calculated for different query size using up to 400 genes using either a spearman correlation (left) or connectivity score (right) approach for drugs perturbed in HEPG2. HMGCR enrichment score was then calculated using the GSEA algorithm. The red dotted line corresponds to a query containing a similar number of up and down regulated genes. Black cross corresponds to a significant enrichment defined as an absolute NES > 1.5. The Black diamond corresponds to the query size when including all significant TWAS genes.

**Supplementary Figure 2:**
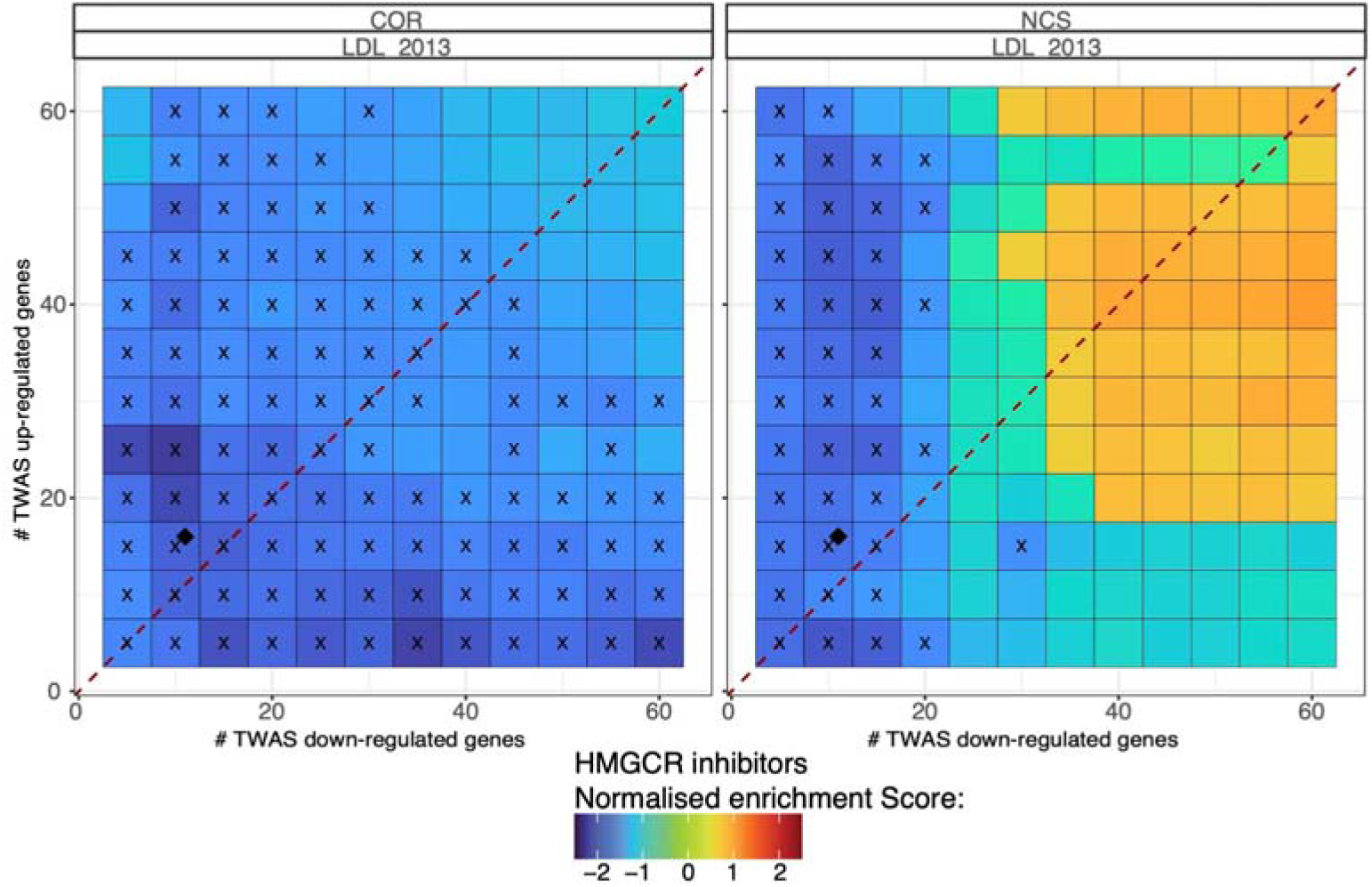
TWAS-based drug repositioning of HMGCR inhibitors for LDL 2013 in HEPG2 cell line using either spearman correlation (left) or normalised connectivity score (right). Normalised enrichment score for HMGCR inhibitors. Individual drug similarity to TWAS signature of a less powered LDL-C GWAS was calculated for different query size using either a spearman correlation (left) or connectivity score (right) approach for drugs perturbed in HEPG2. HMGCR enrichment score was then calculated using the GSEA algorithm. The red dotted line corresponds to a query containing a similar number of up and down regulated genes. Black cross corresponds to a significant enrichment defined as an absolute NES > 1.5. The Black diamond corresponds to the query size when including all significant TWAS genes.

**Supplementary Figure 3:**
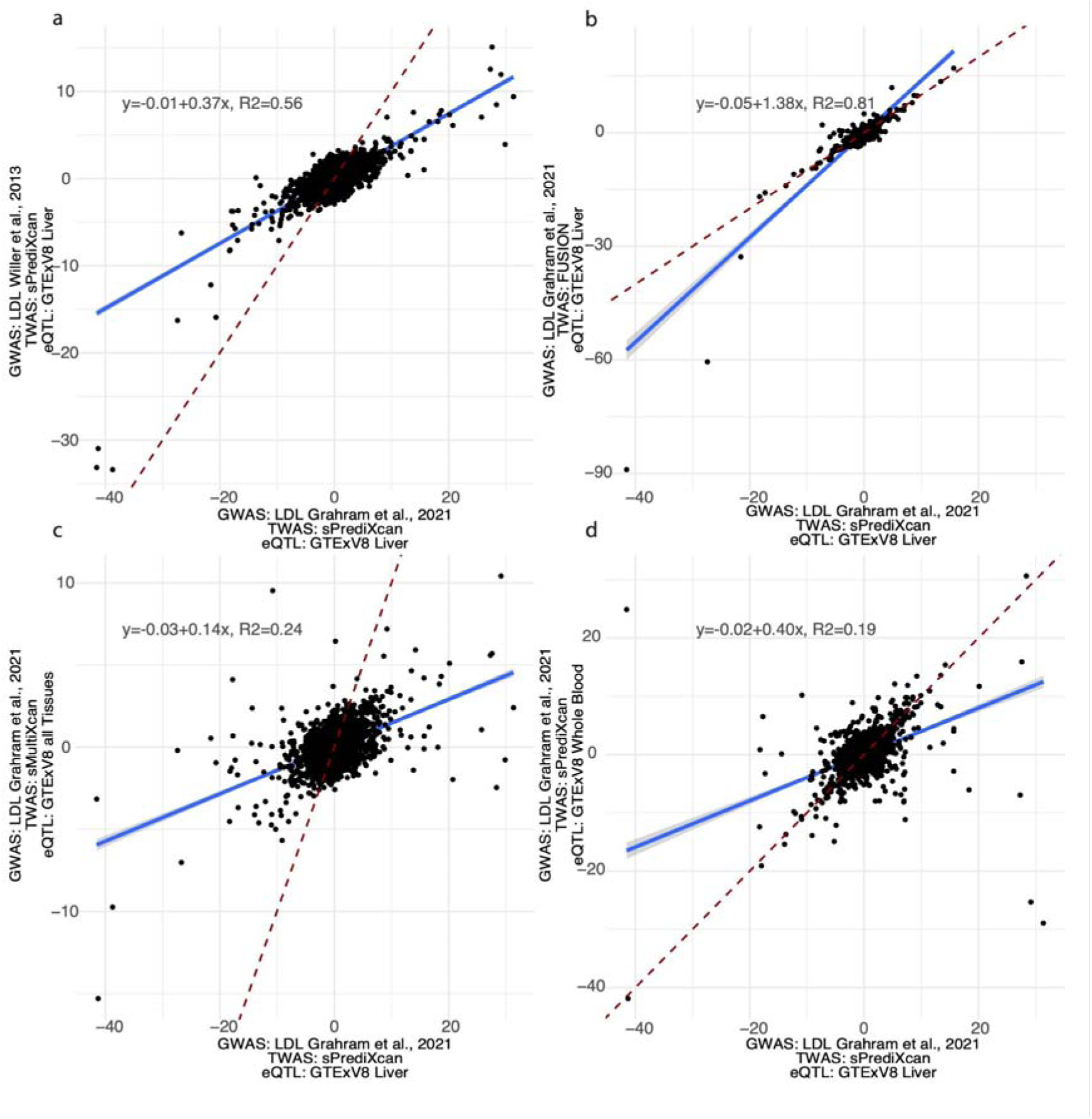
Dot plot showing the correlation between sPrediXcan LDL 2021 GWAS generated using liver eQTL and other LDL GWAS and TWAS methods. a) Dotplot showing the comparison of a TWAS generated using sPrediXcan, liver eQTL and the largest available LDL-C GWAS^29^(Graham et al 2) to a TWAS generated using sPrediXcan, liver eQTL and a less powered LDL-C gwas (Willer et al 2013.,^30^ b) Dotplot showing the comparison of a TWAS generated using sPrediXcan, liver eQTL and the largest available LDL-C GWAS to a TWAS generated using FUSION and the same liver eQTL and GWAS. c) Dotplot showing the comparison of a TWAS generated using sPrediXcan, liver eQTL and the largest available LDL-C GWAS to a TWAS generated usings sMultiXcan with all available GTEx tissues and the same GWAS. d) Dotplot showing the comparison of a TWAS generated using sPrediXcan, liver eQTL and the largest available LDL-C GWAS to a TWAS generated using the same TWAS method and GWAS but with whole blood eQTL.

**Supplementary Figure 4:**
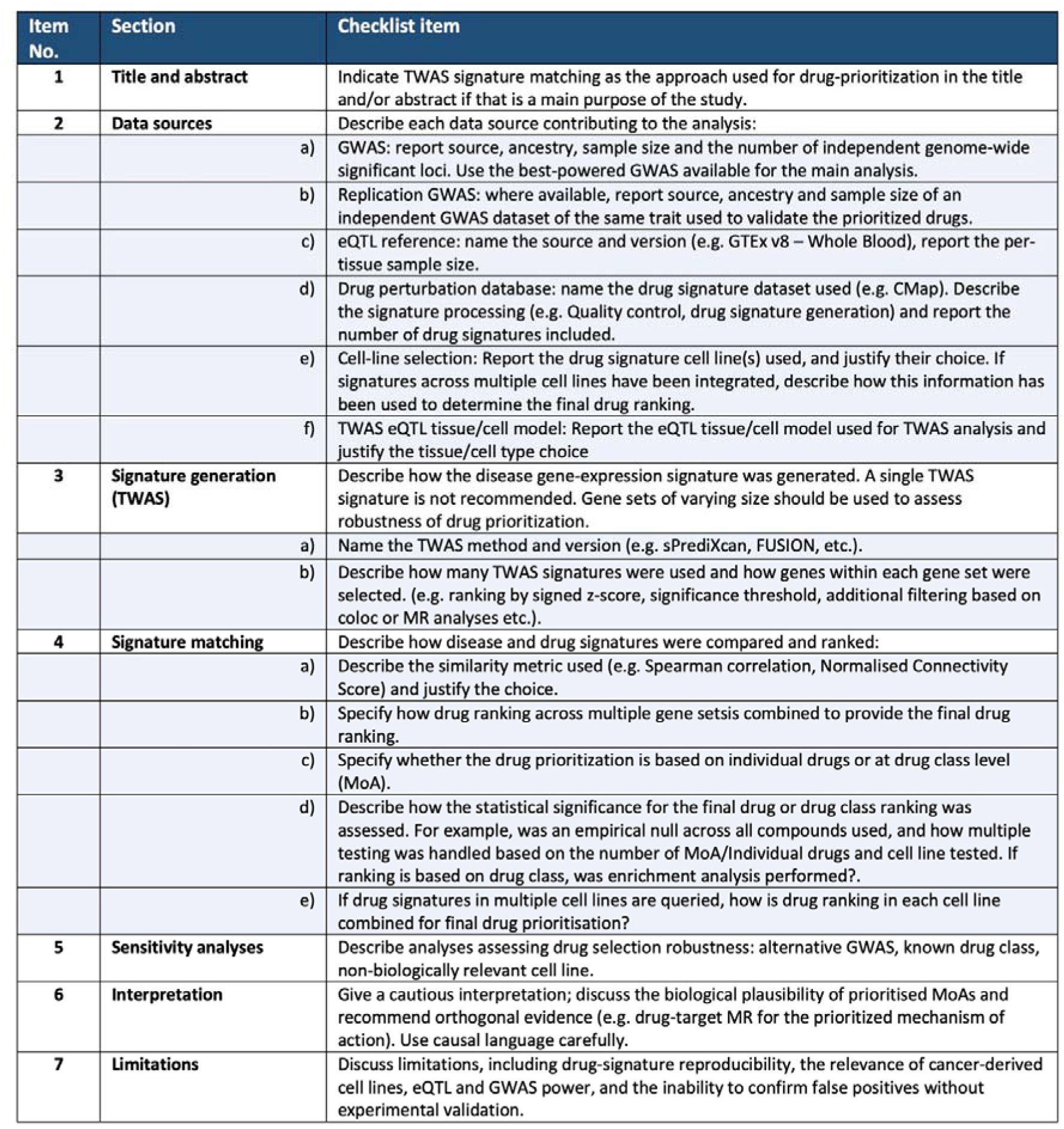
Reporting checklist of recommended items for TWAS signature-matching studies. Checklist of the key methodological decision points and reporting items recommended for transparent and reproducible drug prioritisation using TWAS signature-matching. The checklist accompanies the best-practice framework in Fig. 6 and is intended as a practical guide for authors and reviewers applying this approach.

## Notes

### Competing Interest Statement

The authors have declared no competing interest.

### Funding Statement

This study was funder by ...

### Author Declarations

The study used ONLY openly available human data that were originally located at: Connectivity map (human cell line: https://clue.io/releases/data-dashboard) GWAS summary statistics available from the following studies (PMID ID): 34887591 24097068 34906840 36777996

### Summary of Updates

Fixed typos in the manuscript, as well as added an additional checklist for of recommendation for future TWAS based drug repurposing studies (Supplementary Figure 4 in text)

