## Supplementary Note for "From GWAS to drug: A framework for drug candidate prioritisation using a gene expression signature matching approach"

CMap has been previously reported to have a lack of reproducibility^1^. The focus on profiling a large number of compounds at the expense of a small number of replicates^2^ is a limitation of the study that could lead to a large amount of false positives^3^. Additionally, the overall success rate for the CMap-derived computational predictions is hard to judge leading to uncertainty in the use of CMap. Given this, we investigated the data reproducibility prior to performing TWAS-based drug prioritisation.

#### Data download

We downloaded level 3, 4 and 5 data from the clue.io data dashboard (<https://clue.io/releases/data-dashboard>) (data accessed on 15/06/24). We selected the 2020 version as it combines both phase 1 and phase 2 of the data (Table 1).

###### Table 1. CMap data downloaded from the clue browser.

| **Name** | **Data type** | **Number of signatures** |
| --- | --- | --- |
| Level 3 data | Normalised expression | 1,312,170 |
| Level 4 data | Differentially expressed genes | 1,312,170 |
| Level 5 data | Consensus signature | 510,270 |

#### Compound grouping

We grouped unique compounds based on the following variable: *cmap_name*, *cell_iname*, *pert_itime* and *pert_idose*. This allowed us to identify replicates exposed to the same conditions that could then be grouped together to generate consensus signatures. Untreated (UnTrt) and control wells (DMSO) were removed from the grouping as only active drugs were of interest.

### CMap data distribution

All raw CMap gene expression data was downloaded from the CLUE platform (<https://clue.io/releases/data-dashboard>). Replicate availability for each unique perturbation was assessed across the dataset. We defined a perturbation as a unique combination of drug used (variable name in the dataset: *cmap_name*), cell line where the drug was measurement (*cell_iname*), duration of drug exposure (*pert_itime*), and concentration of the drug during the exposure (*pert_idose*). While looking at the distribution of compounds (Supplementary Note Fig. 1a) prior to generating the consensus signature, we found 151,932 perturbations represented by only a single replicate, and 139,848 by two replicates. We argue that one or two replicates is inadequate to generate a consensus signature and would significantly increase the chances of false positives when shortlisting drugs.

We next evaluated the number of replicates within the consensus signatures (Supplementary Note Fig. 1b). Unexpectedly, we found 105,536 consensus signatures derived from a single replicate and 106,722 from only two replicates despite the implication that Level 5 signatures represent aggregated data. Finally, we observed a large number of duplicated signatures among the 285,223 level 5 signatures based on three replicates, 247,909 were unique while 37,314 were duplicated. Some of the consensus signatures displayed extensive redundancy, for example, cyclosporin A applied to HA1E cells at 10μM for 24 hours had four distinct Level 5 consensus signatures. These findings raise concerns about the consistency of the CMap processing.

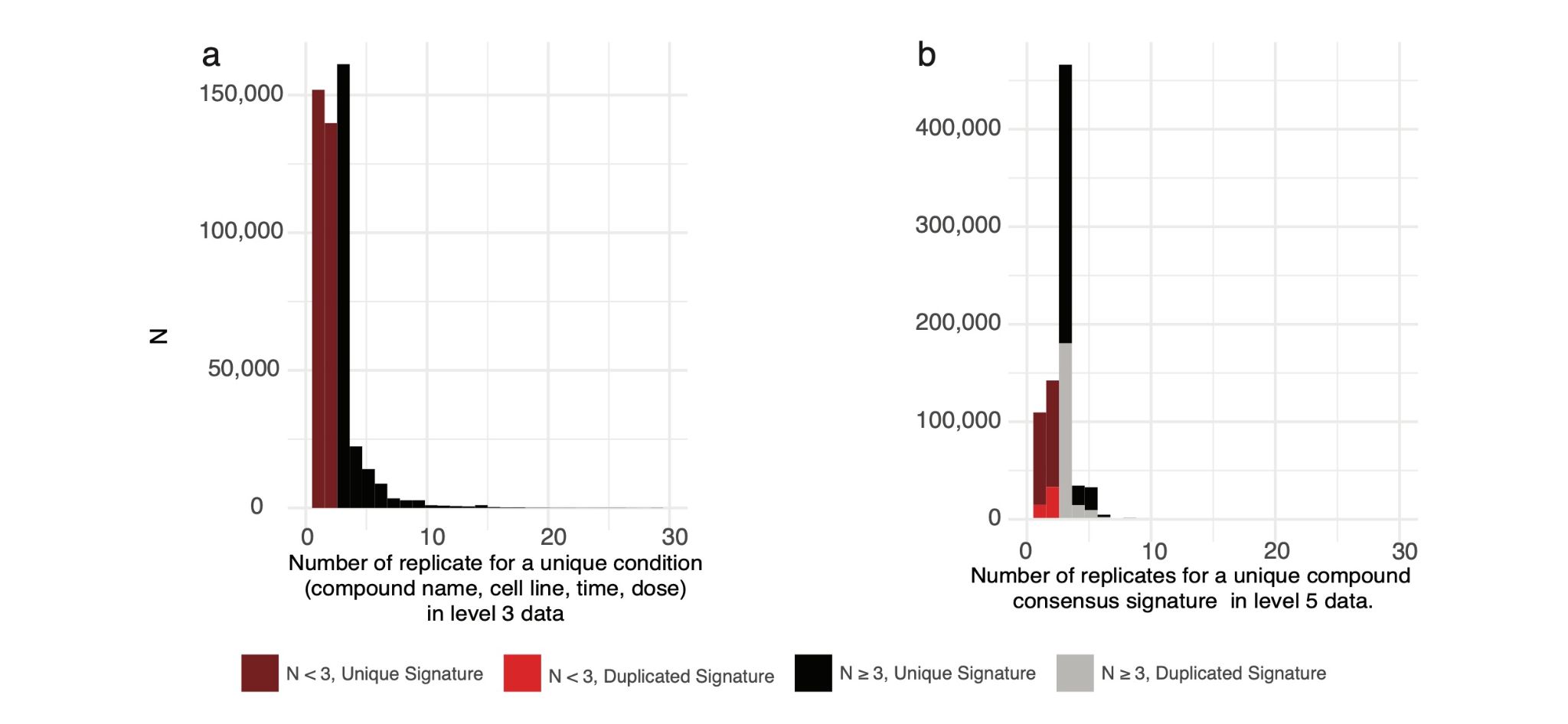

***Supplementary Note Figure 1: CMap data distribution.*** *a. Distribution of the number of replicates found when grouping unique signature compounds based on drug name, cell line, perturbation duration and drug concentration. b. Distribution of the number of replicates used to generate the consensus signatures in the level 5 consensus signatures publicly available from the clue dashboard.*

#### CMAP data reprocessing

The high prevalence of underpowered signatures and the unexpected multiplicity of level 5 profiles led us to reprocess the CMap dataset with the aim of obtaining clean signatures with at least three underlying replicates to perform signature matching approaches.

We started the reprocessing by using Level 3 data (normalised expression) and performed the Level 4 and Level 5 processing following the original Subramanian et al^3^ paper while grouping perturbations as defined previously.

##### L4 normalisation

To obtain a measure of relative gene expression following drug perturbation, Subramanian et al^3^ defined level 4 data as a robust z-score using the following procedure:

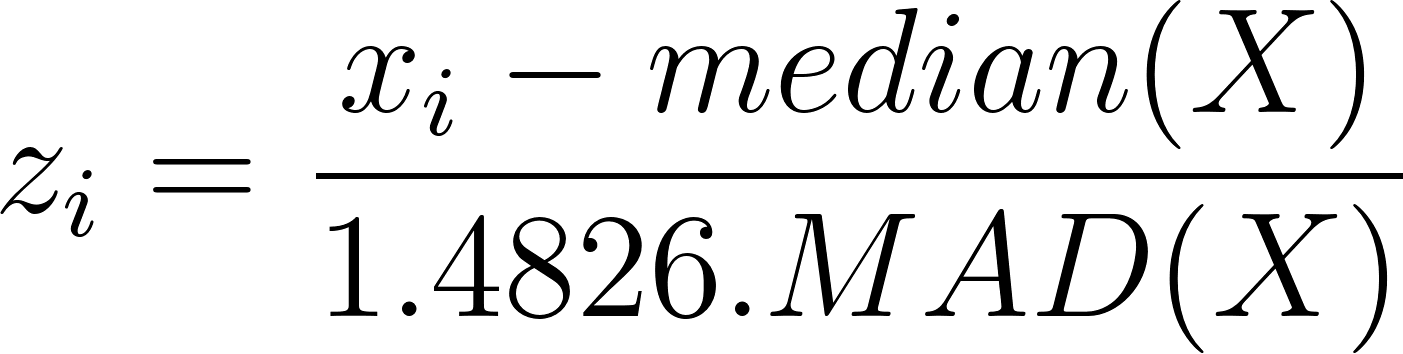
<https://www.codecogs.com/eqnedit.php?latex=z_%7Bi%7D%3D%5Cfrac%7Bx_%7Bi%7D-median(X)%7D%7B1.4826.MAD(X)%7D#0>

Where X is defined as the vector of normalized gene expression (level 3) across all samples on a plate. MAD is the median absolute deviation of X, the factor 1.4826 is used to approximate the standard deviation in the denominator.

##### L5 Signature consensus

Next, we generated consensus signatures, keeping only perturbations with 3 or more replicates.

The consensus signatures were generated by grouping data following:

- Drug used (*cmap_name*)

- Cell type where the measure takes place (*cell_iname*)

- Duration of drug exposure (*pert_itime*)

- Concentration of the drug (*pert_idose*)

We identified 513,747 unique combinations, 151,932 perturbations had only one replicated and 139,848 only 2. Those perturbations were removed leaving a total of 221,967 unique perturbations. The majority (161,250) of those had three underlying replicates, while some such as bortezomib (a compound used as a positive control across conditions) had a large number of replicates. For example, bortezomib measured in the PC3 cell lines for 24 hours at a concentration of 20 uM had 993 replicates.

Following this filtering, we generated Level 5 consensus signatures as follow:

For landmark genes (measured genes), we define the pairwise Spearman correlation matrix between replicates
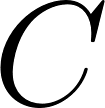
<https://www.codecogs.com/eqnedit.php?latex=C#0> as follow:
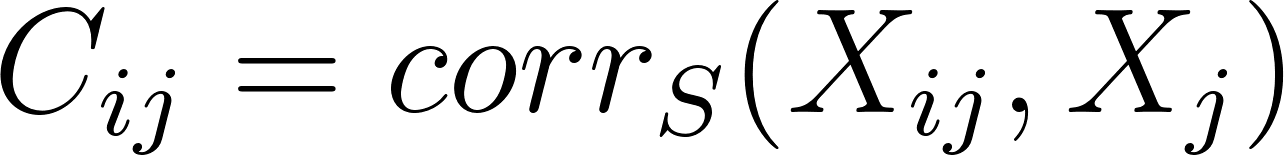
<https://www.codecogs.com/eqnedit.php?latex=C_%7Bij%7D%3Dcorr_%7BS%7D(X_%7Bij%7D%2C%20X_%7Bj%7D)#0>, with self correlations set to zero
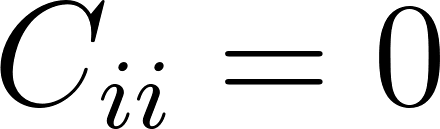
<https://www.codecogs.com/eqnedit.php?latex=C_%7Bii%7D%3D0#0>

The consensus signature is then generated by taking the linear combination of the genes measured within each replicates using the normalised spearman correlation as weight:

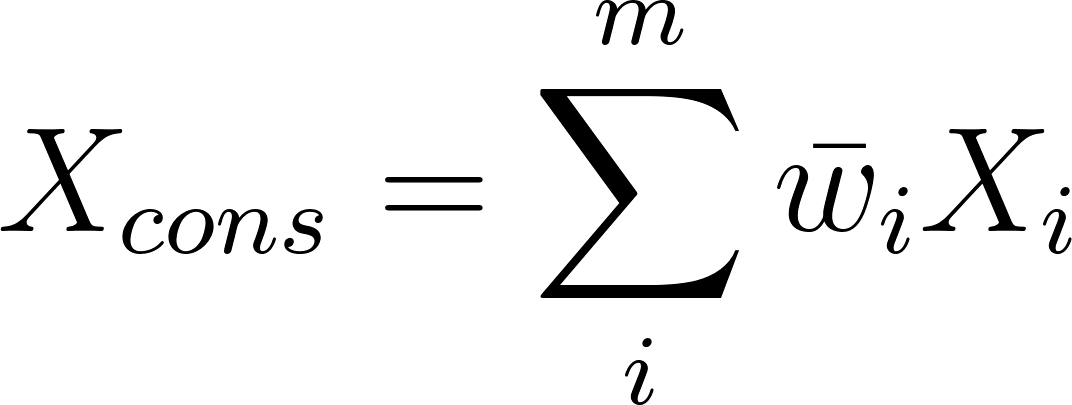
<https://www.codecogs.com/eqnedit.php?latex=X_%7Bcons%7D%3D%5Csum_%7Bi%7D%5Em%5Cbar%7Bw%7D_%7Bi%7DX_%7Bi%7D#0>

The normalisation of the weights for the linear combination is computed as the sum of a replicate correlation to other replicates, normalised to have a sum of 1:

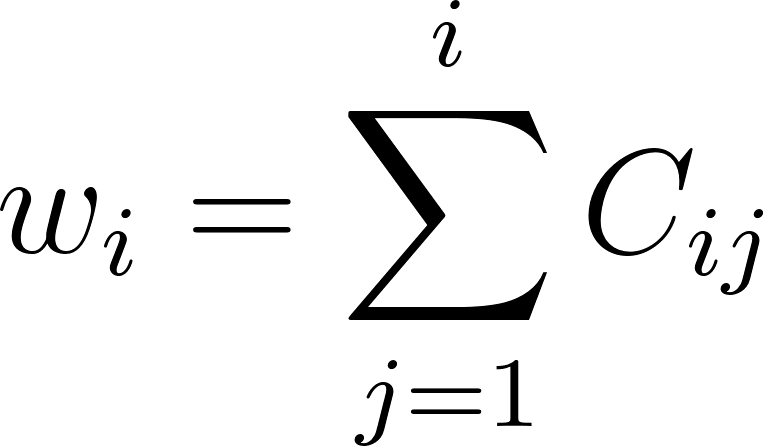
<https://www.codecogs.com/eqnedit.php?latex=w_%7Bi%7D%3D%5Csum%5E%7Bi%7D_%7Bj%3D1%7D%20C_%7Bij%7D%20#0>

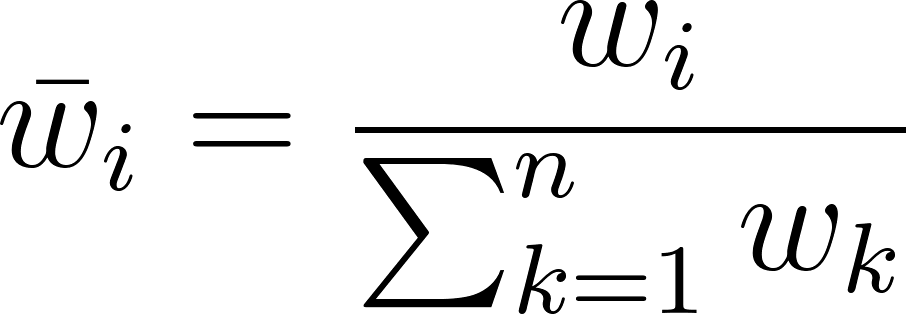
<https://www.codecogs.com/eqnedit.php?latex=%5Cbar%7Bw%7D_%7Bi%7D%3D%5Cfrac%7Bw_%7Bi%7D%7D%7B%5Csum_%7Bk%3D1%7D%5Enw_%7Bk%7D%7D#0>

To illustrate the impact of our reprocessing, we highlighted *HMGA2*, a gene displaying a clear dose-response pattern in HEPG2 cells after 24 hours of exposure when exposed to vorinostat (Supplementary Note Fig. 2a). In the official CMap release, the level 5 data (Supplementary Note Fig. 2c) exhibit multiple level 5 signatures and truncation at a z-score of 10. These artifacts are absent in our reprocessed dataset, while the expected dose-response is preserved, confirming the validity of our reprocessing. Of note, level 4 data (z-score, Supplementary Note Fig. 2b) were identical between our reprocessing and the CMap official release.

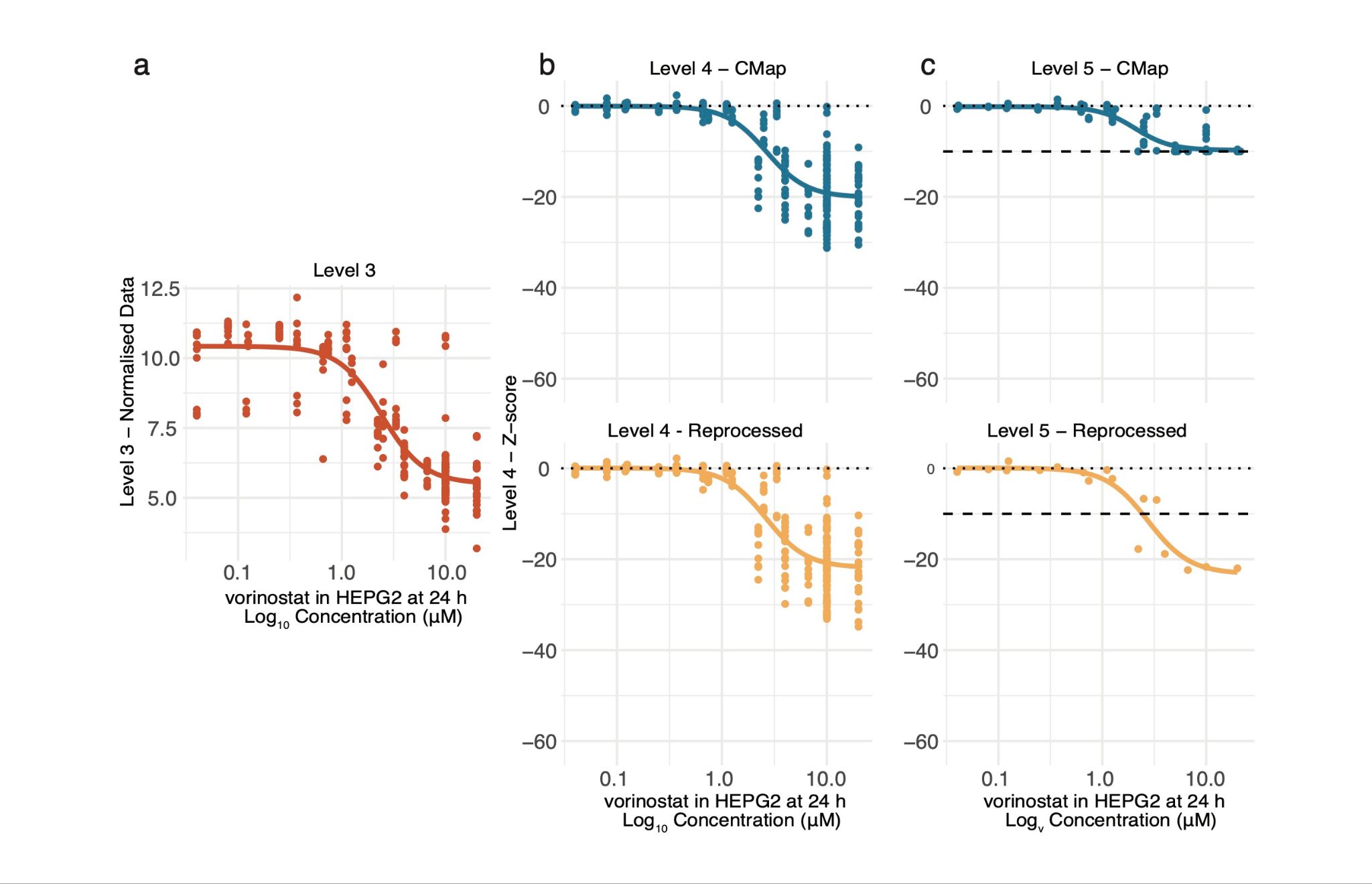

***Supplementary Note Figure 2:*** *CMap reprocessing, example of HMGA2 displaying a dose response when exposed to vorinostat in HEPG2 for 24 hours.
Normalised count (a), z-score processed (b) and consensus signatures (c) of the expression of HMGA2 after administration of vorinostat in HEPG2 for 24 hours is displayed as a function of the concentration used. Blue data labelled CMap corresponds to the already processed data downloaded from the clue portal. Several replicates of the same concentration can be observed at the consensus level. Reprocessed data corresponds to the CMap data reprocessed using the same pipeline as the one highlighted in Subramanian et al.,* ^3^*. The dashed line corresponds to a z-score of -10, the maximum value observed in the downloaded CMap data.*

#### Signature strength:

With the CMap data reprocessed, we next investigated the amount of signal available in the data. Given that the number of genes used to perform signature matching is not defined, we sought to identify the number of perturbed genes when applying different drugs as the upper bound for our signature matching approach. For this, calculated the signature strength (SS) metric referenced in the original CMap paper. SS is defined as the number of differentially expressed genes (DEG) within a drug signature. DEG being a gene displaying a perturbation superior to a threshold of interest.

SS is calculated among landmark genes (measured information) as follow:

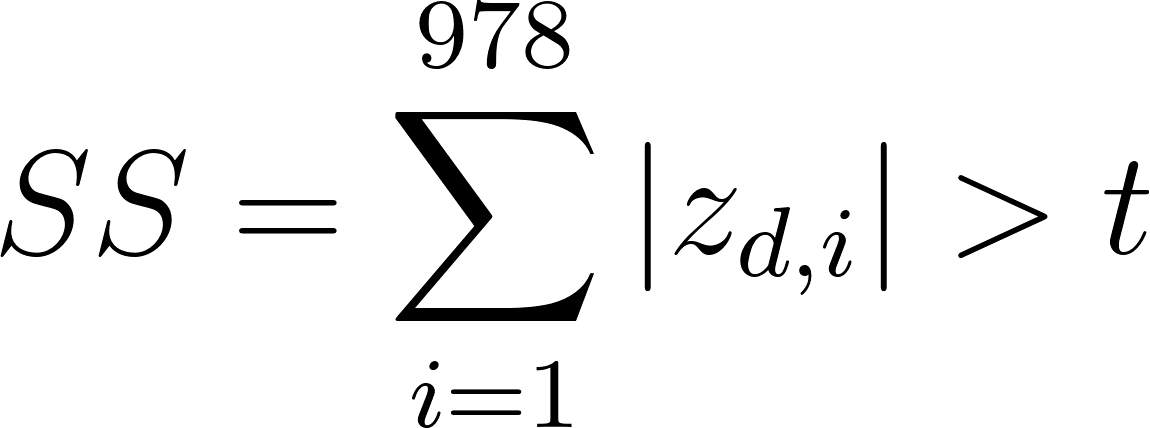
<https://www.codecogs.com/eqnedit.php?latex=SS%3D%5Csum_%7Bi%3D1%7D%5E%7B978%7D%7Cz_%7Bd%2Ci%7D%7C%3Et#0>

With:

-
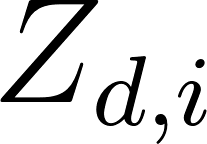
<https://www.codecogs.com/eqnedit.php?latex=Z_%7Bd%2Ci%7D#0> being the value of a consensus signature for a drug
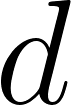
<https://www.codecogs.com/eqnedit.php?latex=d#0> for a specific gene
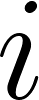
<https://www.codecogs.com/eqnedit.php?latex=i#0>

-
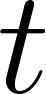
<https://www.codecogs.com/eqnedit.php?latex=t#0> being a threshold of interest.

We varied this threshold
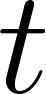
<https://www.codecogs.com/eqnedit.php?latex=t#0> from 0.5 to 4 (Supplementary Note Fig. 3, Supplementary Table 3) and investigated the median number of genes differentially expressed across drugs in CMap with a known mechanism of action. We selected the median instead of the mean as some signatures displayed an extreme number of DEG, biasing the mean upward. We defined DEG as being more than 1.5 standard deviation from the mean (z-score of 1.5) representing value roughly in the top 13% of the distribution. We observed a median of 61 up-regulated and 59 down-regulated genes per consensus signature. Given the limited number of compounds contributing to each signature, as well as the low number of genes passing significance threshold in the consensus signatures; we therefore selected signatures with approximately 60 up- and down-regulated genes as the maximum window for GWAS-based drug repositioning analyses. This number, however, could be refined further to increase the confidence of the DEG selected.

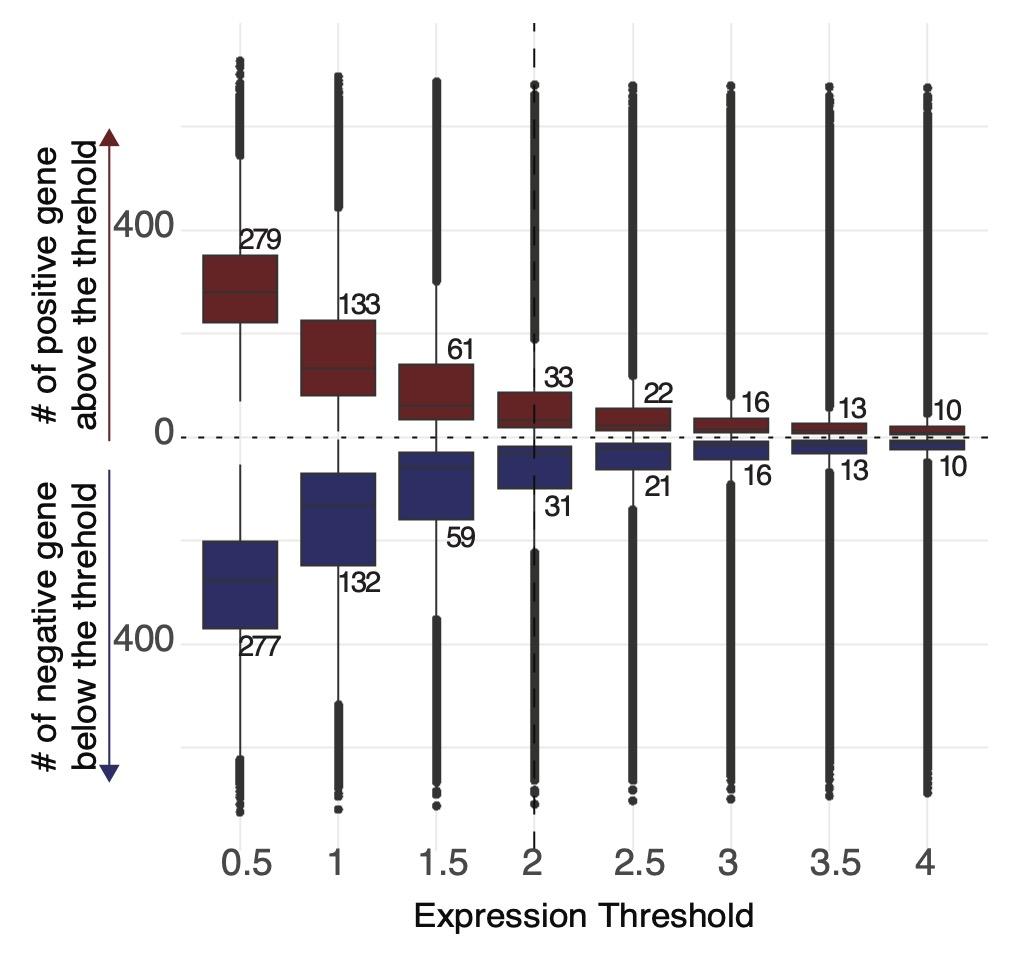

***Supplementary Note Figure 3: Signature strength across CMap.*** *Boxplot representing the number of genes with an absolute value of expression superior to the threshold necessary to be considered as differentially expressed. Numbers written on the boxplot correspond to the median number of DEG positively (red) or negatively (blue) identified.*
